# Mapping social distancing measures to the reproduction number for COVID-19

**DOI:** 10.1101/2020.07.25.20156471

**Authors:** Ellen Brooks-Pollock, Jonathan M. Read, Angela R. McLean, Matt J. Keeling, Leon Danon

**Affiliations:** Bristol Veterinary School, University of Bristol, BS40 5DU; NIHR Health Protection Research Unit in Behavioural Science and Evaluation, Population Health Sciences, Bristol Medical School, University of Bristol, BS8 2BY; Lancaster Medical School, University of Lancaster; Department of Zoology, University of Oxford, OX1 3SZ; Mathematics Institute and School of Life Sciences, University of Warwick, CV4 7AL; CEMPS, University of Exeter; The Alan Turing Institute

**Keywords:** COVID-19 Reproduction number, social contact data, social distancing measures, school closures

## Abstract

**Background:** In the absence of a vaccine, SARS-CoV-2 transmission has been controlled by preventing person-to-person interactions via social distancing measures. In order to re-open parts of society, policy-makers need to consider how combinations of measures will affect transmission and understand the trade-offs between them.

**Methods:** We use age-specific social contact data, together with epidemiological data, to quantify the components of the COVID-19 reproduction number. We estimate the impact of social distancing policies on the reproduction number by turning contacts on and off based on context and age. We focus on the impact of re-opening schools against a background of wider social distancing measures.

**Results:** We demonstrate that pre-collected social contact data can be used to provide a time-varying estimate of the reproduction number (*R*). We find that following lockdown (when *R=*0.7 (95% CI 0.6, 0.8)), opening primary schools as a modest impact on transmission (*R* = 0.89 (95%*CI*: 0.82 − 0.97)) as long as other social interactions are not increased. Opening secondary and primary schools is predicted to have a larger impact (*R* = 1.22, 95%*CI*: 1.02 − 1.53)). Contact tracing and COVID security can be used to mitigate the impact of increased social mixing to some extent, however social distancing measures are still required to control transmission.

**Conclusions:** Our approach has been widely used by policy-makers to project the impact of social distancing measures and assess the trade-offs between them. Effective social distancing, contact tracing and COVID-security are required if all age groups are to return to school while controlling transmission.

## Introduction

The reproduction number, or the ‘R number’ has become a central statistic used to characterise the transmission of severe acute respiratory syndrome–coronavirus 2 (SARS-CoV-2). Early estimates of the reproduction number, which is the average number of secondary cases due to a single case, range between 2.5 and 3.5(1,2), indicating that at least 2 out of every 3 transmission events need to be prevented in order to avoid an outbreak and control an ongoing epidemic. In the United Kingdom (UK), social distancing restrictions, including closing schools, non-essential workplaces, universities, pubs and restaurants, introduced on 23 March 2020, led to an overall reproduction number less than 1 and a decline in the daily number of cases and deaths. The subsequent challenge was to quantify the effect of interventions and their easing on the reproduction number. It is uncertain how the relaxation of these restrictions, especially the physical return to school of the school-age population, will affect the transmission of the virus, though contact tracing and isolation of discovered cases is anticipated to mitigate some of the impact.

The reproduction number of close contact infections such as SARS-CoV-2 depends critically on who meets whom. Social contact surveys, which typically ask about an individual’s social contacts on the previous day, are the most direct way of assessing the potential for spread in a population(3,4). Several such surveys have quantified the behaviour of the UK population prior to the pandemic in 2020(5–7); they demonstrated strong age-assortative mixing patterns and an average number of contacts per person around 12. Surveys conducted in the UK during the COVID-19 pandemic have shown that social distancing dramatically decreased the average number of social interactions to less than 3 contacts per person per day(8).

Social distancing measures, such as the closure of schools and workplaces and mandatory reduction of social interactions, while effective at preventing transmission, have severe economic and psychological effects, and of particular concern is their impact on children(9). Age-specific behavioural patterns mean that social distancing measures affect age groups differently. In normal circumstances, the majority of social contact hours for persons over 60 years of age occur at home while only a quarter of their social contact hours are associated with leisure activities outside the home. In contrast, nearly 60% of twenty to thirty-year olds’ social contact hours are at work(6). Crucially, nearly half of children’s social contact hours are made within a school setting, meaning that school closures have a major impact on the social experience of young people. In this study, we use social contact data(6), including an additional targeted survey of children, to quantify the impact of re-opening schools on the reproduction number in the UK(10).

## Materials and Methods

### Social Contact Data

We used data from the Social Contact Survey (SCS) which surveyed 5,861 individuals in the UK in 2010 about their social contacts during a single day(6). Participants were recruited using three approaches: a paper survey sent to people in the post, an online survey and an online survey aimed specifically at school-aged children. Participants were asked to complete demographic information about themselves including age, occupation and about their social contacts on the previous day. Participants were asked to report the number of people they met, the duration of the contact (<10 minutes, 10 to 59 minutes, 1 to 4 hours, 4+ hours), the context (home, work/school, travel, other/leisure), and whether the contact involved touch, e.g. a handshake, hug or kiss. To facilitate reporting large numbers of contacts per day, participants could report contacts as individual contacts or groups of contacts; this methodology better captures the right-hand tail of the degree distribution. Participants were also asked about transitive interactions between contacts, for more details see(11).

### Estimating the Reproduction Number from social contact data

We use an individual-based approach to calculate a reproduction number of each of the participants of the SCS study(10). The reproduction number for an individual is generated as a sum of all social interactions, multiplied by the probability of transmission given the interaction:

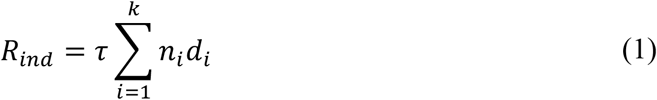

where *k* is the number of contact events reported by each participant, *n*_*i*_ is the number of individuals in that contact (groups of similar contacts), *d*_*i*_ is the duration of the contact and *τ* is the probability of transmission. Because we do not have ages of contacts, this is an ego-centric estimate of R, and does not include local depletion of susceptibles.

The population-wide reproduction number, *R*_*t*_, represents the *average* number of secondary cases due to an *average* infectious person. As both the risk of becoming infected and the risk of infecting others is proportional to the number of contacts, individuals with more contacts will contribute more to *R*_*t*_ than individuals with fewer contacts. Therefore *R*_*t*_ will depend on the sum of the squared individual reproduction numbers, i.e.

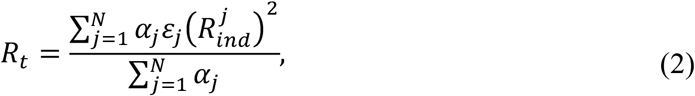

where *N* is the number of participants in the Social Contact Survey, 0 ≤ *ε*_*i*_ ≤ 1 is the relative infectiousness of children under 11 years of age relative to adults. *α*_*i*_ is the age-specific weighting for participant *j*, estimated to match the age distribution of the UK population, calculated as the ratio of the proportion of individuals aged *a* in the UK, *P*_*UK*_(*a*), to the SCS sample, *P*_*SCS*_(*a*),

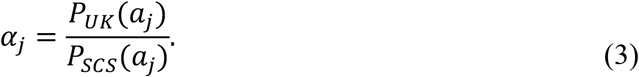

### Model calibration (estimating τ)

The model can be calibrated using incidence data when the social contact patterns are known. Here, we calibrated the model to the exponential growth phase of the epidemic in the UK prior to the introduction of widespread social distancing on 23 March 2020. We estimated the growth rate, *g*, from death data between 13 March 2020 and 30 March 2020, then calculated the reproduction number as *R* = exp (*gS*) where *S* is the serial interval.

### Estimating the reproduction number following stay-at-home order on 23 March 2020

Google has made community mobility reports(12) available for the period during COVID-19 transmission from 15 February 2020. The Google mobility reports provide a point estimate for the percentage change in number of visits to, and length of stay at places categorised as grocery and pharmacy, parks, transit stations, retail and recreation, residential, and workplace. The median percentage change is relative to the median value for the same day of the week for the period between 3 January 2020 and 6 February 2020(12).

We mapped the context reported in the SCS onto the Google mobility data categories as home is equivalent to residential, work/school to workplace, other/leisure to retail and recreation and travel to transit. We assumed that 100% of contacts were active during the week of 18 March 2020. We then used the Google mobility estimate of the percentage of contacts that were active in subsequent weeks.

### Forward simulating social distancing measures and school closures

#### 1. Generalised social distancing measures

To simulate *x*% of contacts in a given context being active, we take a random sample without replacement of a proportion (1 − *x*/100) of all contacts for that context. The selected contacts are flagged with a comply flag *c*_*i*_ equal to 1. The reduced individual reproduction number is given by:

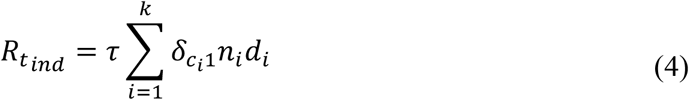

Where 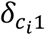 equals zero if *c*_*i*_ = 1 and one otherwise.

#### 2. School closures

Primary and secondary schools were closed in the UK from 23 March 2020. It was estimated that approximately 2% of pupils attended school during April and May 2020 as children of essential workers(13). To simulate school closures, we remove all contacts for the relevant school aged children that have “school” listed as the context by setting *d*_*i*_ = 0. To capture the 2% of children who remained at school, we re-instated a random sample of the removed contacts. We do not capture additional contacts due to school, such as parent-to-parent contact.

#### 3. “COVID security”

COVID security is a term used to describe the reduction in transmission due to the wearing of face coverings, eye protection and maintaining physical distancing during social interactions(14). In the UK, face coverings became mandatory on public transport and NHS settings in June 2020, and in shops and supermarkets from 24 July 2020. We capture COVID security by reducing the transmission probability *τ* by 25% and 50%.

#### 4. Contact tracing

We modelled contact tracing as implemented in the UK, that is, triggered by a symptomatic test-positive case. For each individual, we assign a probability of symptoms given infection based on their age. We used the age-specific symptomatic rates estimated in (15): cases under 18 years of age have a 25% chance of symptoms given infection, cases over 80 years of age have a 75% chance of symptoms given infection and we impose a linear increase with age between the two ages(15). For each individual, we draw a random number to determine if they are symptomatic and eligible for contact tracing. We assume that contact tracing has the effect of reducing their number of secondary infections by the contact tracing efficacy; we consider example scenarios where 20% and 60% of contacts are successfully traced.

For each of the *R*_*t*_ estimates, we calculate the mean and 95% confidence intervals for the reproduction number by sampling contacts then bootstrapping contacts, weighted by age, 2000 times and taking the percentile confidence interval.

### Data and code availability

Social Contact Survey data are available at http://wrap.warwick.ac.uk/54273/. Data, code and example implementation are available at github.com/ellen-is/reckoners.

## Results

### Model calibration and baseline values

We estimate that the number of deaths in the UK grew exponentially with a rate of 0.23 (95% CI 0.22, 0.24) deaths per day between 13 March 2020 and 30 March 2020. This corresponds to a reproduction number of 2.7 using a mean serial interval of 7.5 days (16–18).

We combine this estimate of the reproduction number prior to lockdown with social contact data to estimate a transmission probability per contact hour of 0.002 hour^-1^, see Materials and Methods for interpretation of this value.

### Impact of the UK stay-at-home order on 23 March 2020

Following lockdown, we use Google Community Mobility Reports as a proxy for the percentage reduction in active work, leisure and travel contacts. With a 65% reduction in work contacts, a 75% reduction in leisure contacts and a 95% reduction in school contacts, the reproduction number is reduced to 0.7 (95%CI 0.6, 0.8) (figure 1), which is consistent with direct estimates from social contact surveys(8).

**Figure 1:**
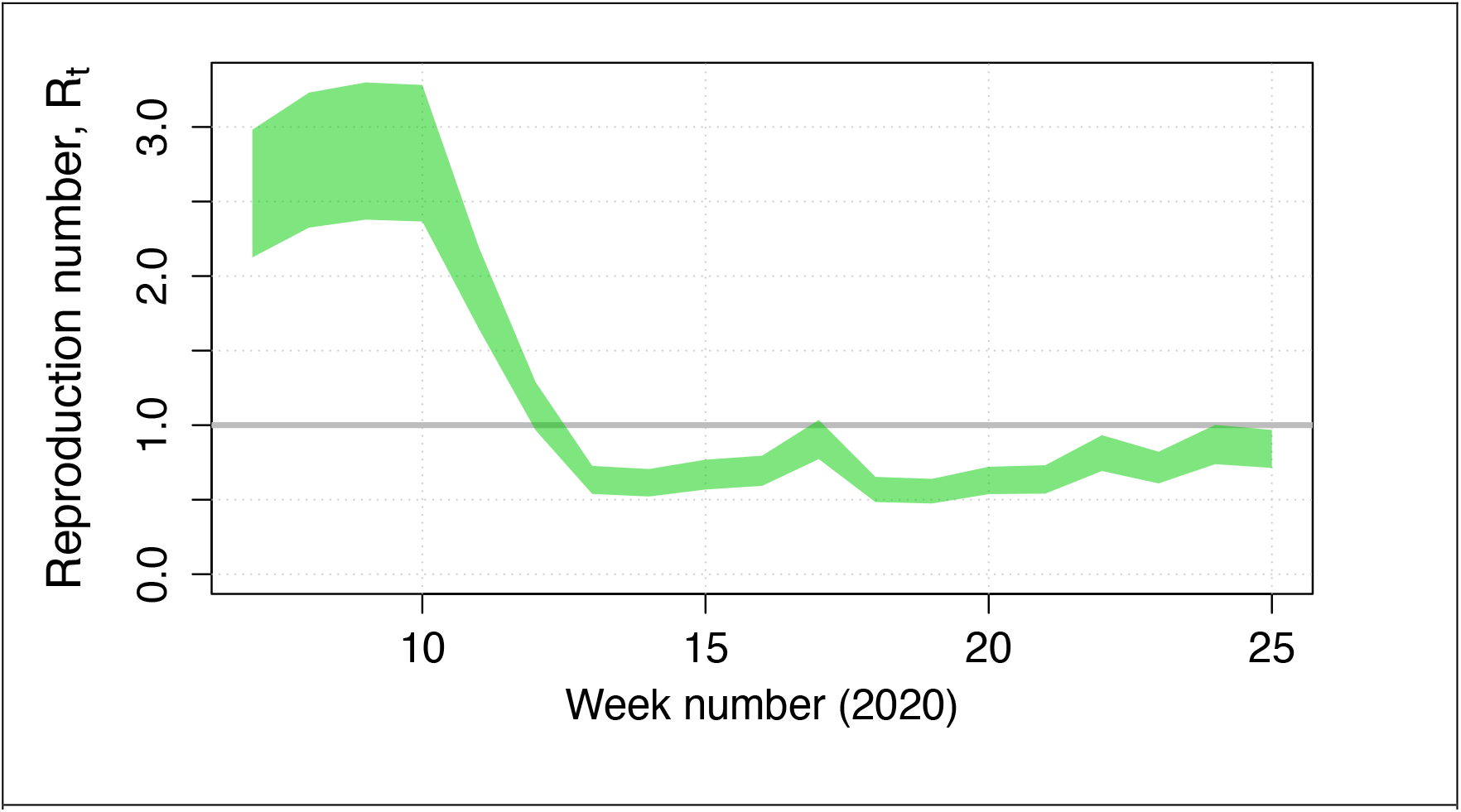
The time-varying reproduction number in the UK, estimated using incidence death data prior to lockdown, Social Contact Survey data and Google Community Mobility Reports.

### Contribution of social contacts by context

We assessed of the relative importance of social contacts by context. Preventing all leisure and other contacts, while allowing work, school and household contacts, leads to a reproduction number of 2.0 (95%*CI*: 1.8 − 2.4). Preventing work contacts while allowing leisure and household contacts has a bigger impact, resulting in a reproduction number of 1.5 (95%*CI*: 1.4 − 1.7). Using this approach, the minimum reproduction number without preventing household contacts is 0.45 (95% CI, 0.41, 0.50). We stress that this estimate does not allow for essential contacts outside the home due to keyworkers and essential services but provides a lower bound for the reproduction number in the UK.

### Impact of contact tracing

Tracing and isolating contacts of symptomatic cases can be effective at reducing the reproduction number, however extremely high levels of contact tracing need to be achieved for the reproduction number to be brought close to 1 without social distancing measures (figure 2). As expected, the more contacts traced per case, the more effective contact tracing is at reducing the reproduction number.

**Figure 2:**
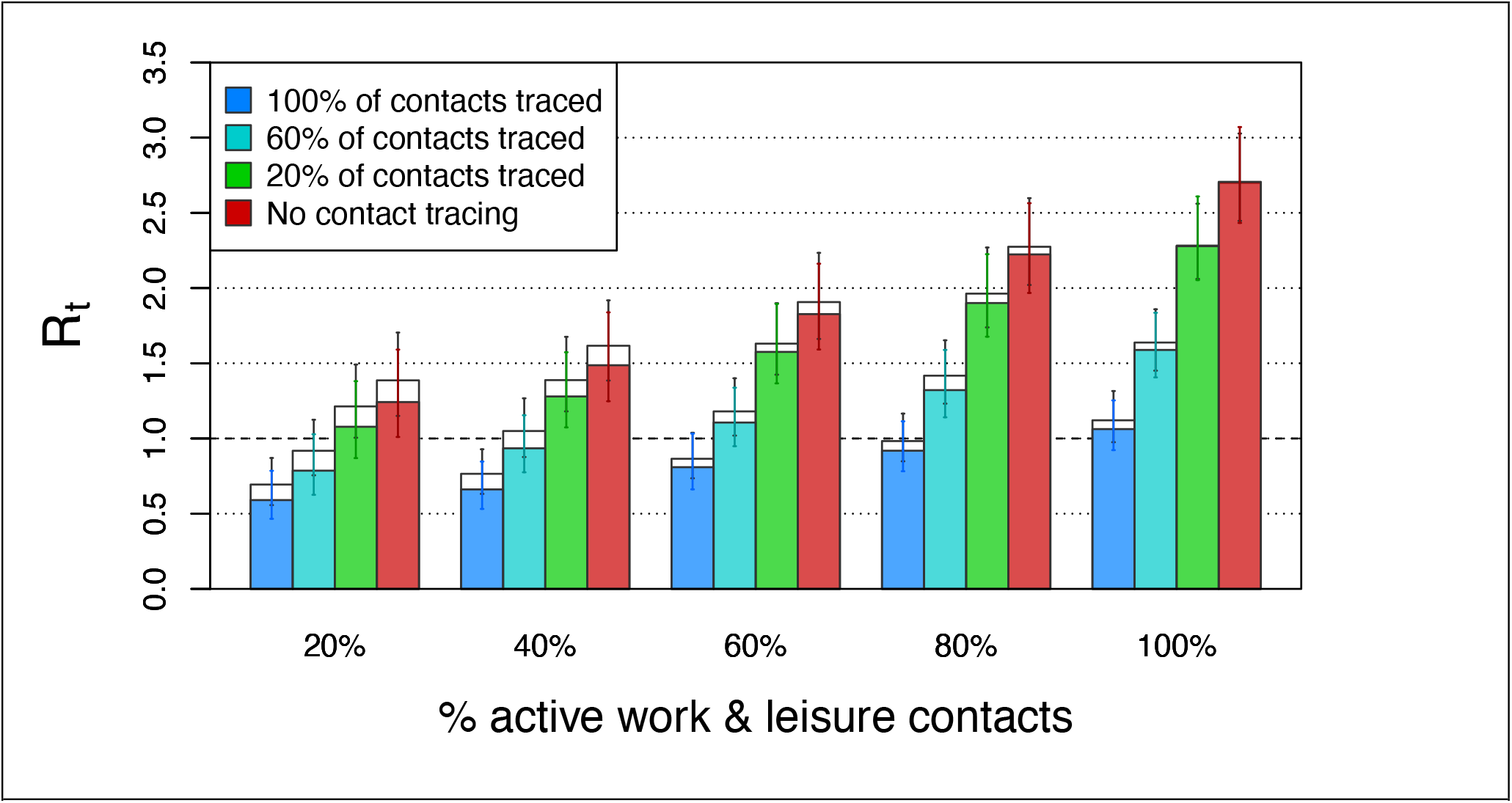
The impact of tracing and isolating contacts of symptomatic cases when all children are back at school. The empty bars are under the assumption that children under 11 years of age are as infectous as adults; the filled bars are where children under 11 years of age are half as infectious as adults.

The absolute effectiveness of contact tracing is dependent on concurrent social distancing measures. With strict social distancing similar to levels shortly after the start of the stay-at-home order, contact tracing is able to prevent up to ∼0.7 secondary infections per case. In contrast, with no social distancing, contact tracing can prevent up to ∼1.6 secondary infections per case. Assumptions about the infectiousness of children have a minimal impact on these conclusions (figure 2).

### The impact of multiple interventions and school closures

Figures 3a-3i show the projected reproduction number for school re-opening scenarios as a function of the percentage of pre-COVID active social contacts and varying levels of contact tracing and COVID security.

**Figure 3:**
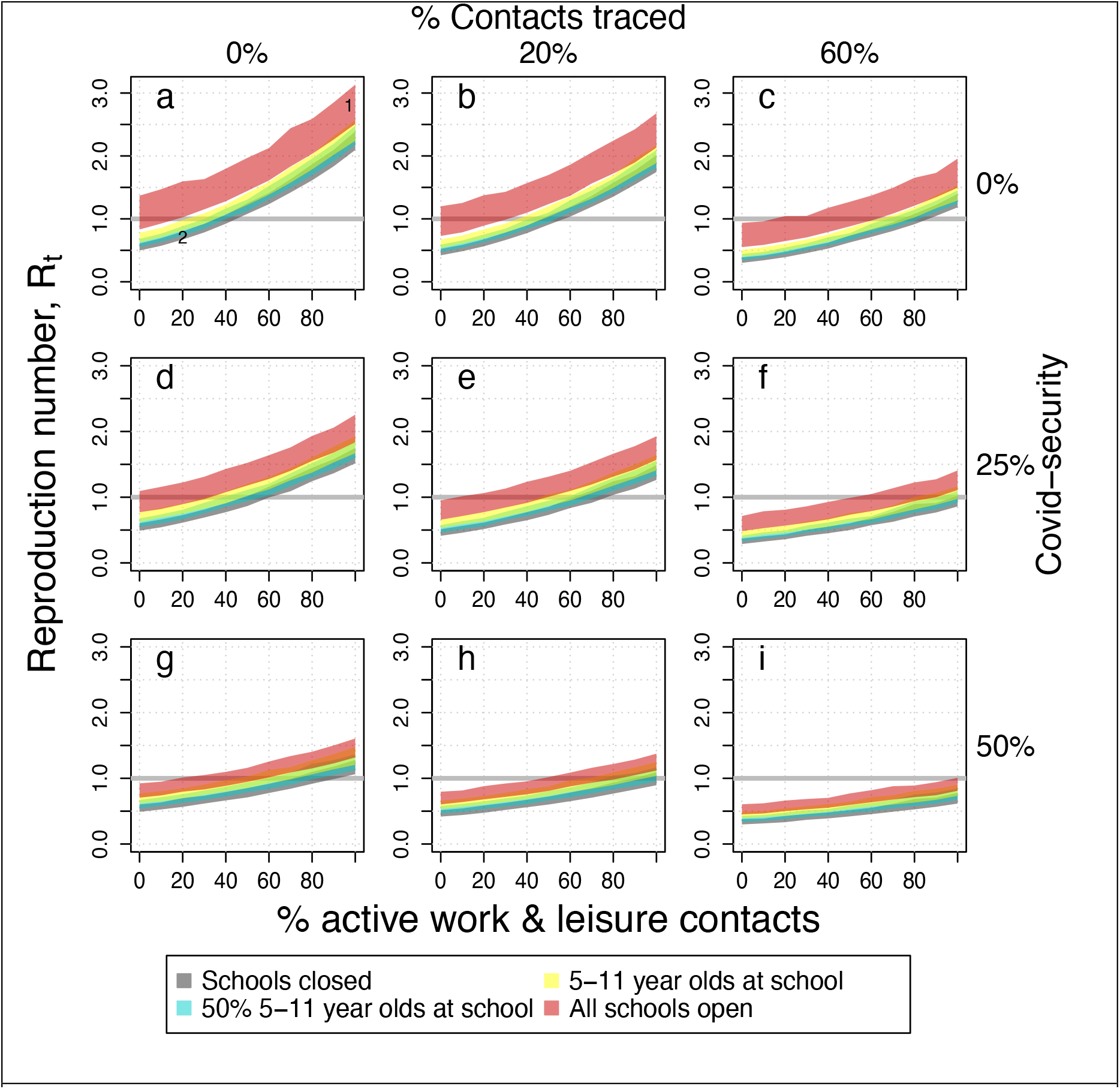
The COVID-19 reproduction number as a function of the percentage of active work and leisure contacts under different contact tracing, COVID-security and school closure scenarios. The shaded regions indicate the values with schools closed (grey), 50% of primary school pupils attending school (cyan), all primary school pupils at school (yellow) and primary and secondary schools open (red). The panels illustrate the values of the reproduction number with (a) no contact tracing or COVID-security and with increasing measures (b-i). The number 1 marks the UK position in March 2020, number 2 marks the UK position in April/May 2020. The width of the ribbons indicates 95% confidence intervals. These figures are generated with the assumption that children under 11 years of age are as infectious as adults.

Figure 3a represents no contact tracing or COVID-security. The reproduction number in March 2020, with schools fully open, is marked with the number 1, and the reproduction number in May 2020 with schools closed to all but children of essential workers is marked with the number 2.

Policy options can be mapped to the reproduction number by traversing across the figure panels. For example, from point 2 with schools closed, social distancing measures could be eased so that 40% of pre-COVID contacts occur while maintaining a reproduction number less than 1. However, then, in order to reopen schools while controlling transmission, 60% effective contact tracing would have to be introduced (figure 3c) and/or increased COVID-security (figures 3f-3i).

We find that if no other social contacts outside the home increase apart from those occurring within primary schools, then re-opening primary schools is consistent with a reproduction number less than 1, *R* = 0.89 (95%*CI*: 0.82 − 0.97) (figure 3a). However, even a modest increase in contacts outside home and school, relative to post-lockdown levels, would push the reproduction number back above 1. In the absence of substantial population-level immunity, the additional opening of secondary schools is likely to result in sustained transmission in the population (*R* = 1.22, 95%*CI*: 1.02 − 1.53). In general, higher adherence to other social distancing measures is required as more children return to school.

We predict that contact tracing and increased COVID-security could increase the options for opening schools (figures 3b-i). We assume that a given proportion of all contacts are successfully traced, self-isolate, and that their contribution to the reproduction number is effectively zero. Under a scenario similar to the situation in May/June 2020, where 20% of contacts were effectively traced and isolated, all pupils could return to school if COVID-security could halve transmission and 40% of other contacts were prevented (figure 3h).

However, if 60% of contacts of symptomatic cases were traced and isolated, we estimate that schools could fully re-open while maintaining control of transmission, if at least 60% of other contacts are prevented (*R* = 0.92, 95%*CI*: 0.78 − 1.1) with no COVID -security. In this scenario, other forms of social distancing, including working from home and eliminating leisure contacts, would still be required if schools were to be fully open before a pharmaceutical solution is found. If 50% COVID-security could be achieved in combination with 60% contact tracing, then potentially low levels of social distancing might be required (figure 3i).

Repeating the analysis under the assumption that children are less infectious than adults yields similar conclusions. If children are less infectious than adults then re-opening primary and secondary schools has a smaller impact on the reproduction number, but the impact of increasing other contacts outside home and school settings remains the same.

## Discussion

In this paper, we present a method for mapping social distancing policies to a reproduction number for COVID-19 using social contact data. Focussing on school closures and their subsequent re-opening, we explore the balance between social distancing, contact tracing and COVID-security that is required to control transmission. Our findings suggest that high adherence to social distancing outside school settings is needed to maintain epidemic control without effective contact tracing and/or COVID security. Opening primary schools has a modest impact on R, while opening secondary schools is predicted to have a larger overall impact, but that keeping all schools open should be feasible with a combination of other measures.

Our findings support the use of contact tracing as a key part of epidemic control; however, tracing needs to be highly effective. After the introduction of Test, Trace and Isolate in the UK only 20% of social contacts of cases were successfully traced and isolated within 48 hours, though this has substantially increased over time(19). While tracing 20% of contacts has a positive impact on the reproduction number, it is insufficient to prevent epidemic growth if all schools are fully open.

The greater risk arises from contact with people outside the home and school contexts. It is likely that reopening of schools will also lead to an increase in contacts made outside school, due to caregivers returning to work and interactions between parents. A strength of this analysis is its predictive value regarding the effect of combined interventions. Using metrics of adherence to social distancing measures, such as Google mobility or contemporary social contact surveys, it is possible to map a country’s progression out of lockdown, and therefore estimate the effect of policy changes on the reproduction number (20).

Other studies have used social contact data to characterise the impact of social distancing measures on COVID-19 transmission. Retrospective estimates of the reproduction number have been mapped to social distancing policies post implementation to estimate the relative effectiveness of measures(21,22). In these analyses, re-opening schools was associated with an increase in the reproduction number, although it is not possible to discriminate between social contacts in education and other settings. The limited data on young people’s contact patterns makes simulating the impact of school closures challenging. Simulations which included a fixed increase in work and community contacts as a result of schools re-opening found that the associated increase in incidence could be mitigated by effective contact tracing and additional testing(23,24) and that school closures on their own were not sufficient reduce the reproduction number to below unity(25), in line with our conclusions

The limited understanding of young people’s contact patterns is one of the limitations of our analysis, and our approach does not include some of the complexities and non-linearities observed in disease dynamics. The Social Contact Survey data that we used are built up around disconnected “egos”, so our approach does not capture household structures, clusters, cliques and higher-level social organisation which influence epidemic spread at a population level. Furthermore, as the epidemic progresses, immunity plays an increasingly important role in dynamics. Our approach uses the basic reproduction number to characterise transmission, and therefore does not capture the build-up of immunity in a population as all contacts are assumed to be susceptible to infection. Depending on the age distribution of immunity, social distancing measures are likely to lead to different changes in the reproduction number.

This analysis was made possible by pre-existing detailed social contact data. Social contact patterns have been used to characterise the potential for disease transmission in a population(26), design vaccine and control programmes for infectious diseases including influenza(27), meningitis(28) and now COVID-19(8). However, in many settings, such data are out-of-date or not available. Given their proven value, we argue that regular, representative social contact surveys should be become a routine part of epidemic control and preparedness.

## Data Availability

All data used for the paper are publicly available.

http://wrap.warwick.ac.uk/54273/

## Acknowledgement and funding statement

The authors gratefully acknowledge comments and discussions from the members of the Scientific Pandemic Influenza Group on Modelling (SPI-M) for useful comments and discussions. This work was partly supported by the National Institute for Health Research Health Protection Research Unit (NIHR HPRU) in Evaluation of Interventions at the University of Bristol (EBP), NIHR grant MEMVIEER NIHR200411 (MJK), The Alan Turing Institute EPSRC EP/N510129/1 (LD), Medical Research Council grant MC/PC/19067 (LD, EBP, MJK), MR/V038613/1 (MJK, EBP, LD, JMR), MR/5004793/1 (JMR) and Engineering and Physical Sciences Research Council grant EP/N014499/1 (JMR).

